# Pharmacotherapy for adults with metabolic dysfunction-associated steatotic liver disease (MASLD) and metabolic dysfunction–associated steatohepatitis (MASH): a systematic review and network meta-analysis

**DOI:** 10.1101/2025.09.03.25335039

**Authors:** Yousef Manialawy, Erin Deacon, Téa Sue, Ramsha Khan, Vihaan Sharma, Joseph Maasarani, Areeb Jafrani, Ariya Chumber, Jeremy Steen, Rachel Couban, Matthew Collins, Puneeta Tandon, Mang Ma, Kailei Nong, Xinyu Zou, Hang Sun, Yongfeng Song, Sheyu Li, João PL Lima, Behnam Sadeghirad, Gordon Guyatt, Arnav Agarwal

**Author notes:** **Correspondence to:** Arnav Agarwal, Division of General Internal Medicine, University of Alberta, 5-112 Clinical Sciences Building, 11304-83 Ave NW, Edmonton, AB, Canada, T6G 2B7. Shared first authorship (Yousef Manialawy, Erin Deacon).

## Abstract

**Introduction:** Metabolic dysfunction-associated steatotic liver disease (MASLD) and metabolic dysfunction-associated steatohepatitis (MASH) have risen substantially in prevalence over recent decades, driven by a growing global burden of obesity, diabetes mellitus, and other cardiometabolic risk factors. In response, researchers have intensified efforts to evaluate novel and re-purposed therapies that may prevent or reverse disease progression. Although several pharmacological therapies are under investigation, robust comparative evidence on their relative effectiveness and safety remains limited. We will therefore conduct a systematic review and network meta-analysis (SRNMA) of randomized controlled trials (RCTs) evaluating pharmacological therapies for adults with MASLD or MASH.

**Methods:** We will search four electronic databases (Ovid MEDLINE, Embase, CINAHL and Cochrane CENTRAL) from inception to August 2025 without language and other restrictions. Eligible studies will include parallel-arm RCTs enrolling ≥10 adults per arm with MASLD or MASH; comparing any pharmacological therapy to standard care, no treatment, lifestyle modifications, placebo or alternative pharmacotherapies; and having a minimum follow-up duration of 12 weeks. Primary clinical outcomes are all-cause mortality, cardiovascular mortality, hospitalization, progression to cirrhosis, hepatic decompensation, hepatocellular carcinoma, and serious treatment-related adverse events. Surrogate outcomes include histological, imaging, biochemical, and metabolic markers of disease activity.

Paired reviewers will independently screen identified hits for eligibility, extract data from eligible studies, and assess risk of bias using the Risk Of Bias instrument for Use in SysTematic reviews-for Randomised Controlled Trials (ROBUST-RCT). We will conduct separate NMAs for MASLD and MASH populations using a frequentist graph-theoretic random-effects model. Certainty of evidence will be assessed using GRADE. Subgroup and sensitivity analyses will explore effect modification by comorbidities and study quality.

**Ethics and Dissemination:** No ethics approval is required. Results will be disseminated via peer-reviewed publication and conference presentations to inform clinicians, guideline developers, and health system decision-makers.

**PROSPERO Registration Number:** CRD420251103235.

## INTRODUCTION

Metabolic dysfunction-associated steatotic liver disease (MASLD) affects nearly 30% of the global population and has a rising global prevalence, in line with increasing prevalences of associated cardiometabolic risk factors including obesity, type 2 diabetes, dyslipidemia and hypertension.^1–5^ MASLD can progress to metabolic dysfunction-associated steatohepatitis (MASH), characterized by hepatic inflammation and fibrosis, and can ultimately lead to cirrhosis with complications such as ascites, variceal bleeding and hepatocellular carcinoma.^6–8^ MASLD is now the second leading indication for liver transplantation worldwide, reflecting both its rising prevalence and clinical burden.^9,10^

Management of MASLD and MASH currently emphasize modification of metabolic risk factors, reducing further liver impairment through measures such as alcohol abstention, hepatitis A and B vaccination, and active management of metabolic comorbidities.^11–16^ Pharmacological options have chronically been limited to select therapies such as vitamin E, pioglitazone in patients with diabetes, and more recently to the THR-β agonist resmetirom.^17–19^ There remains a need for additional pharmacotherapies to improve the prognosis of adults with MASLD and MASH, and for reliable evidence regarding their impact on patient-important outcomes. In response, new pharmacotherapies are rapidly emerging, and numerous trials evaluating existing and emerging therapies are being conducted.^20^

A comprehensive evidence-based summary evaluating the comparative effectiveness and safety of available pharmacotherapies for MASLD and MASH is essential to inform the decisions of clinicians, patients, guideline developers and decision-makers in healthcare systems. Network meta-analysis, rather than pairwise meta-analysis, provides the optimal method to determine the relative effects of treatments that have not been tested head-to-head. This systematic review and network meta-analysis therefore addresses the identified gap by evaluating the effectiveness and safety of pharmacotherapies for adults with MASLD and MASH.

## METHODS AND ANALYSIS

The protocol for this systematic review adheres to Preferred Reporting Items for Systematic Reviews and Meta-Analyses (PRISMA)-Protocols standards. We will adhere to the PRISMA checklist for reporting our systematic review and meta-analysis. Our protocol is also registered on PROSPERO (CRD420251103235).

### Search strategy

With the help of a medical librarian, we will conduct a systematic search of the published literature in four electronic databases from inception to present: Ovid MEDLINE (1946– August 2025), Embase (1947– August 2025), CINAHL (1982– August 2025) and the Cochrane Central Register of Controlled Trials (1991– August 2025).

Search terms will include a combination of keywords and Medical Subject Heading (MeSH) terms, including “randomized controlled trial”, “metabolic dysfunction-associated steatotic liver disease”, “MASLD”, “metabolic dysfunction-associated steatohepatitis”, “MASH”, “non-alcoholic fatty liver disease”, “NAFLD”, “non-alcoholic steatohepatitis”, “NASH”, “metabolic dysfunction-associated fatty liver disease”, “MAFLD”, “fatty liver” and individual drug names and classes.

We will also supplement our search by reviewing the reference lists of related systematic reviews and included studies trials (i.e. backward searching). We will not apply language or other restrictions in our literature search. The full search strategy is included as an Appendix.

### Eligibility criteria

Eligible studies will be parallel-arm randomized controlled trials (RCTs) including adults diagnosed with imaging or biopsy-confirmed MASLD, or elastography or biopsy-confirmed MASH, and reporting on at least one pre-specified outcome of interest. Studies with participants with steatotic liver disease caused exclusively by known secondary etiologies other than metabolic-related disease including viral hepatitis, alcoholic hepatitis, autoimmune hepatitis, or drug induced hepatitis will be excluded. Participants with cirrhosis at study onset, regardless of etiology, will be excluded.

Eligible interventions include any pharmacological therapy compared to standard care, no treatment, lifestyle modifications, placebo, or alternative pharmacotherapies aimed at treating MASLD or MASH. Where clinically appropriate, treatments will be classified based on drug class or therapeutic groups. Studies systematically comparing combinations of pharmacological therapy where arms differ in more than one treatment group will be eligible. Studies exclusively comparing different doses of the same treatment will be ineligible.

Herbal products, botanical products, phytomedicines and traditional medicines will be ineligible. We will exclude studies and interventions that risk violating joint randomizability (i.e. transitivity assumption).

We will include unpublished trials with available data via trial registries or other grey literature sources. We will not apply language or other restrictions. Eligible trials will have a minimum follow-up of 12 weeks and randomize at least 10 participants per arm. We will exclude quasi-randomized and cross-over trials.

Clinical outcomes of interest are:

1. All-cause death.
2. Cardiovascular death.
3. Admission to hospital.
4. Progression to cirrhosis confirmed on imaging or biopsy.
5. Hepatic decompensation, including gastrointestinal bleeding, varices, ascites, hepatic encephalopathy, spontaneous bacterial peritonitis, hepatopulmonary syndrome, hepatorenal syndrome, hepatocellular carcinoma, related splenomegaly.
6. Serious treatment-related adverse events.

Surrogate outcomes of interest are:

1. Histological improvement on liver biopsy defined as a reduction in steatosis, steatohepatitis, lobular inflammation, hepatocellular ballooning, and/or fibrosis grade. Improvement can be reported using any standardized scoring tool.
2. Imaging-based improvement and/or resolution of steatosis.
3. Imaging-based improvement and/or resolution of fibrosis using elastography-derived liver stiffness measurement.
4. Change in liver enzymes (particularly alanine aminotransferase levels).
5. Change in non-invasive risk estimation scores (e.g. FIB-4).
6. Change in metabolic markers (glycated hemoglobin, body weight, body mass index, markers of insulin resistance or glucose intolerance, lipid panel).

For all clinical outcomes other than adverse events, we will extract data if reported at a minimum follow-up duration of 24 weeks. Outcome data will be extracted at the timepoint closest to 24 weeks (± 4 weeks) and at the latest reported point of follow-up. For adverse events and surrogate outcomes, we will extract data at a minimum follow-up duration of 12 weeks (and up to 16 weeks), closest to 24 weeks (± 4 weeks) and at the latest reported point of follow-up. Trial-defined definitions will be used across outcomes unless otherwise specified.

### Study selection

Using pre-tested standardized screening forms with accompanying instructions and following completion of a pilot exercise, paired reviewers will screen identified hits at title-and-abstract and full-text levels. Reviewers will resolve disagreements by discussion, with adjudication by a third reviewer when necessary. We will use Covidence (https://covidence.org/) for screening and study selection. Reasons for exclusion at the full-text screening level will be summarized using a PRISMA flow diagram.

### Data extraction and risk of bias assessment

Paired reviewers will independently extract data and assess risk of bias for each included study at the outcome level using pre-tested, standardized extraction forms with accompanying guidance. Discrepancies will be resolved through discussion and, if necessary, adjudication by a third reviewer.

We will extract data related to the study population, interventions, comparators, and outcomes of interest. For the population, the following variables will be collected: (1) study characteristics (year, countries, setting, funding, number of patients, length of follow-up); (2) patient characteristics (sex, age, race, diagnosis of MASLD/MASH including method of diagnosis, severity of MASLD/MASH per standardized criteria used in the trial, diagnosis of metabolic risk factors as previously defined, and relevant complications as previously defined); (3) interventions (drug name, class, dose, duration); (4) outcome data (outcomes, definitions, follow-up durations; for dichotomous outcomes, number of events, number of patients analyzed; for continuous outcomes, mean or median values and measures of statistical variability).

Paired reviewers will independently assess risk of bias (RoB) via the ROBUST-RCT tool, which assesses bias across the following domains: random sequence generation (selection bias), allocation concealment (selection bias), blinding of participants and personnel (performance bias), blinding of outcome assessment (detection bias), incomplete outcome data (attrition bias), selective outcome reporting (reporting bias), and other potential sources of bias (e.g., early trial termination, influence of funding).^21^ Each domain is evaluated using a series of signaling questions, with one of four judgements possible: ‘‘low’’, ‘‘probably low’’, ‘probably high’’, or ‘‘high’’. For each domain, we will evaluate whether methodological safeguards are present, and whether concerns regarding risk of bias exist related to the trial methods.

### Assessment of certainty of evidence

We will assess the overall certainty of evidence regarding each direct, indirect and network estimate using the Grading of Recommendations Assessment, Development and Evaluation (GRADE) approach. We will use either the null or outcome-specific minimal important differences (MID) as the decision threshold of our rating. We will rate certainty for every direct comparison based on RoB, inconsistency (heterogeneity), indirectness, and publication bias, and will categorize overall certainty as high, moderate, low, or very low, starting at high for RCTs. A contribution matrix will quantify the proportional contribution of each direct comparison to each network estimate using the random-walk approach, which will determine the initial certainty for indirect and network estimates. The certainty of indirect estimates may be further downgraded due to intransitivity. We will determine certainty of network estimates accounting for incoherence (between direct and indirect estimates) and imprecision (wide CI). Ratings for precision will consider whether 95% CI cross the decision threshold, and to account for the possibility of inflated effect sizes from small bodies of evidence, whether the optimal information size (OIS) is met for large effect sizes.

### Statistical analysis

We will conduct separate network meta-analyses for studies evaluating adults with MASLD and those evaluating adults with MASH.

For binary outcomes, we will pool effect estimates using risk ratios (RR) with 95% confidence intervals (CI). To facilitate interpretation, we will calculate absolute risk differences by applying the pooled RR to the baseline risk for each outcome. Baseline risks will be estimated using event rates from the control arms of included studies. For continuous outcomes, we will pool effect estimates using mean differences (MD) with 95% CI and will use change scores from baseline. Continuous outcomes reported using different measures or scales will be transformed to a common scale using natural units of the most frequently reported instrument or scale across included studies.

We will employ a frequentist graph-theoretic framework for random-effects network meta-analysis, implementing weighted least squares estimation with Moore-Penrose pseudoinverse computation. Analyses will be performed using the netmeta package in R software.

We will consider grouping treatments into common nodes based on pharmacological class or treatment group. Groupings will not account for duration or dose of treatment. Expert gastroenterologists and methodologists will be consulted to verify appropriateness of categorizations and definitions used for treatment nodes.

We will evaluate for the global coherence (consistency) assumption using a design-based decomposition of Cochran’ Q statistic; a p-value of less than 0.05 will be interpreted as significant global inconsistency (i.e. consistency assumption is rejected at the overall level of each treatment). Direct estimates will be calculated by conducting pairwise meta-analysis for all comparisons with direct evidence in the network. We will evaluate between-trial heterogeneity using the I^2^ statistic and visual assessment of forest plots; I^2^ values between 0% and 40% will be interpreted as likely unimportant heterogeneity, 30% to 60% as likely moderate heterogeneity, 50% to 90% as likely substantial heterogeneity, and 75% to 100% as considerable heterogeneity. Where at least 10 trials contribute to a comparison, we will evaluate for small study effects through visual inspection of funnel plots and Harbord’s score test for binary outcomes or Egger’s method for continuous outcomes; where funnel plot asymmetry is observed or the results of the test show statistically significant small study effects, we will perform a sensitivity analysis excluding small studies. Indirect estimates will be calculated using node splitting and back calculation method. We will evaluate for comparison-specific incoherence by considering clinical and statistical differences between direct and indirect estimates. Where incoherence is observed, we will investigate for possible sources including errors in extracted data.

We will estimate ranking probabilities among treatments using the surface under the cumulative ranking curve (SUCRA), mean ranks and rankograms.

If sufficient data are available, we will plan the following subgroup analyses for pairwise meta-analyses and meta-regression for network meta-analyses to examine heterogeneity in treatment effects:

- Presence versus absence of concomitant type 2 diabetes (hypothesis: larger treatment effects in patients with concomitant type 2 diabetes).
- Presence versus absence of concomitant cardiovascular disease (hypothesis: larger effects in patients with established cardiovascular disease, particularly for mortality, cardiovascular and kidney outcomes).
- Presence versus absence of concomitant chronic kidney disease (hypothesis: larger effects in patients with established chronic kidney disease, particularly for mortality, cardiovascular and kidney outcomes).
- Presence versus absence of concomitant obesity (hypothesis: larger effects in patients with obesity).

We will perform a sensitivity analysis limiting the analysis to studies at low risk of bias.

We will rate the credibility of effect modification analyses using the Instrument for assessing the Credibility of Effect Modification Analyses (ICEMAN) tool.^18^

## Data Availability

All data included in the study will be available upon reasonable request to the authors once the study is formally published.

## ETHICS AND DISSEMINATION

As this study is a systematic review and network meta-analysis of previously published data, ethics approval is not required. We intend to disseminate our findings through presentations at national and international medical conferences. In addition, we plan to submit the final manuscript for publication in a peer-reviewed journal that is widely read by general practitioners, internists, and other healthcare professionals concerned with liver disease.

## PATIENT AND PUBLIC INVOLVEMENT

The research question and study design were developed with a focus on patient-important outcomes. Patients and members of the public will not be directly involved in the design of this review.

